# Emergence of two distinct SARS-CoV-2 Gamma variants and the rapid spread of P.1-like-II SARS-CoV-2 during the second wave of COVID-19 in Santa Catarina, Southern Brazil

**DOI:** 10.1101/2022.01.13.22268697

**Authors:** Dayane A. Padilha, Vilmar Benetti-Filho, Renato S. Moreira, Tatiany A. T. Soratto, Guilherme A. Maia, Ana P. Christoff, Fernando H. Barazzetti, Marcos A. Schörner, Fernanda L. Ferrari, Carolina L. Martins, Eric K. Kawagoe, Julia K. Wachter, Paula Sacchet, Antuani R. Baptistella, Aline D. Schlindwein, Bruna K. Coelho, Sandra B. Fernandes, Darcita B. Rovaris, Marlei P. D. Anjos, Fernanda R. Melo, Bianca Bittencourt, Sthefani Cunha, Karine L. Meneghetti, Nestor Wendt, Tamela Z. Madaloz, Marcus V. D. Rodrigues, Doris S. M. Souza, Milene H. Moraes, Rodrigo P. Baptista, Guilherme Toledo-Silva, Guilherme R. Maciel, Edmundo C. Grisard, Patrícia H. Stoco, Luiz F. V. Oliveira, Maria L. Bazzo, Gislaine Fongaro, Glauber Wagner

## Abstract

The Western mesoregion, the State of Santa Catarina (SC), Southern Brazil, was heavily affected as a whole by the COVID-19 pandemic in early 2021. This study aimed to evaluate the dynamics of the SARS-CoV-2 virus spreading patterns in the SC state through March 2020 to April 2021 using genomic surveillance. During this period, 23 distinct variants, including Beta and Gamma, among which, the Gamma and related lineages were predominant in the second pandemic wave within SC. A regionalization of P.1-like-II in the Western-SC region was observed, concomitant to the increase in cases, mortality, and case fatality rate (CFR) index. This is the first evidence of the regionalization of the SARS-CoV-2 in SC transmission and highlights the importance of tracking variants, dispersion, and impact of SARS-CoV-2 on the public health systems.

## Introduction

The COVID-19 pandemic, caused by the SARS-CoV-2 virus, is the world’s most significant public health challenge of the last 100 years. This virus has infected more than 270 million people, leading to more than 5,3 million deaths worldwide by December 2021 [*1*]. In the same period, since the first reported case on 12/01/2019, this virus infected around 22.2 million people leading to 617 thousand deaths in Brazil and in the Southern State of Santa Catarina (SC), 1,238,056 cases and 20,101 deaths were reported [*2*].

With the virus’ rapid accumulation of polymorphisms and the advance of the pandemic new variants have emerged [*3*]. Thus, genomic surveillance has been essential for monitoring this disease and vital for local health agencies to have information to mitigate the effects of the pandemic and adopt strategies to increase population engagement in COVID control policies [*4*].

Until December 2021, more than 6,1 million SARS-CoV-2 genomes have been sequenced, allowing detection of several variants [*5*]. Some new variants have distinctive biological characteristics from their predecessors, such as better transmission, immunological escape or distinguishing clinical symptoms [6]. Some variants of SARS-CoV-2 have been defined as Variants of Interest (VOIs) or Variants of Concern (VOCs) with potential impact on public health. The main VOCs described have been B.1.1.7/VOC 202012 (Alpha), December 2020; B.1.351/501Y.V2 (Beta), December 2020; P.1/ B.1.1.248/B1.1.28 (Gamma), January 2021; and B.1.617.2 (Delta), May 2021 [*1*]. Recently, the B.1.1.529 (Omicron) variant was reported in South Africa in November 2021 [1], evidencing that new VOC of SARS-CoV-2 are still emerging and, thus, genomic surveillance of the virus remains crucial.

In Brazil, the VOC Gamma (P.1) variant was first reported in the city of Manaus, State of Amazonas, in November 2020, being identified in 42% of RT-PCR positive samples collected between December 15^th^ and 23^th^, 2020 [*7*]. This VOC has 1.7-2.6 times more transmissible than previous strains circulating in Brazil, showing a 1.2-1.9-fold increase in the mortality risk to adults and 5-8-fold for the young population [*8*], broadening the susceptible population under risk of infection and hospitalization and the needs of hospitalization and intensive care [*9*].

Among several mutations for VOC Gamma the E484K mutation occurs in the receptor binding domain (RBD) domain of the S protein and is believed to assist the evasion of the virus from the immune system [*10*] and, therefore, may lead to lower vaccine efficacy [*11*]. Also present in the RBD domain, the N501Y mutation is associated with increased binding specificity to the host cell angiotensin-converting enzyme 2 (ACE2) receptor. Subvariants of VOC Gamma and related lineages (P.1-like-I and P.1-like-II), displaying distinct mutations, have also emerged during 2021 [*12*]. Among these, P.1.1, P.1.2, and P.1-like-II have spread across the border States of Rio Grande do Sul [*13*] and Paraná [14] up to April 2021 and, concomitantly detected in SC state [*12*].

In this context, our study evaluated the dispersion of different variants of SARS-CoV-2 through March 1^st^, 2020 to April 30^th^, 2021, in the state of Santa Catarina, especially during the second wave of the pandemic (January to April 2021) when the VOC Gamma variant became predominant in Brazil.

## Material and methods

### Studied regions and sampling

The State of Santa Catarina (SC) is located in the southern region of Brazil, having borders with the Paraná (North) and Rio Grande do Sul (South) states, and Argentina (West). With an estimated population of 7,333,473 inhabitants, the SC state is geographically divided into six mesoregions: Greater Florianópolis, Itajaí River Valley, Mountain range, Northern, Southern, and Western SC [*15*].

Genome sequences from 203 SARS-CoV-2 positive samples collected from different mesoregions of the state of Santa Catarina, Brazil through March 1^st^, 2020 to April 30^th^, 2021, were generated (Figure 1) (Supplementary Table 1). Additionally, 527 SARS-CoV-2 genome sequences obtained from samples isolated in SC (Figure 1) and 49 genome sequences from samples isolated worldwide, including the original SARS-CoV-2 sequence (NC_045512.2), were retrieved from the EpiCov database GISAID [*5*] and included in this study (only genome sequences showing <5% ambiguous nucleotide and > 85% genome sequence coverage). This study was approved by the Federal University of Santa Catarina Ethics Committee (CAAE: 31521920.8.0000.0121).

**Figure 1:**
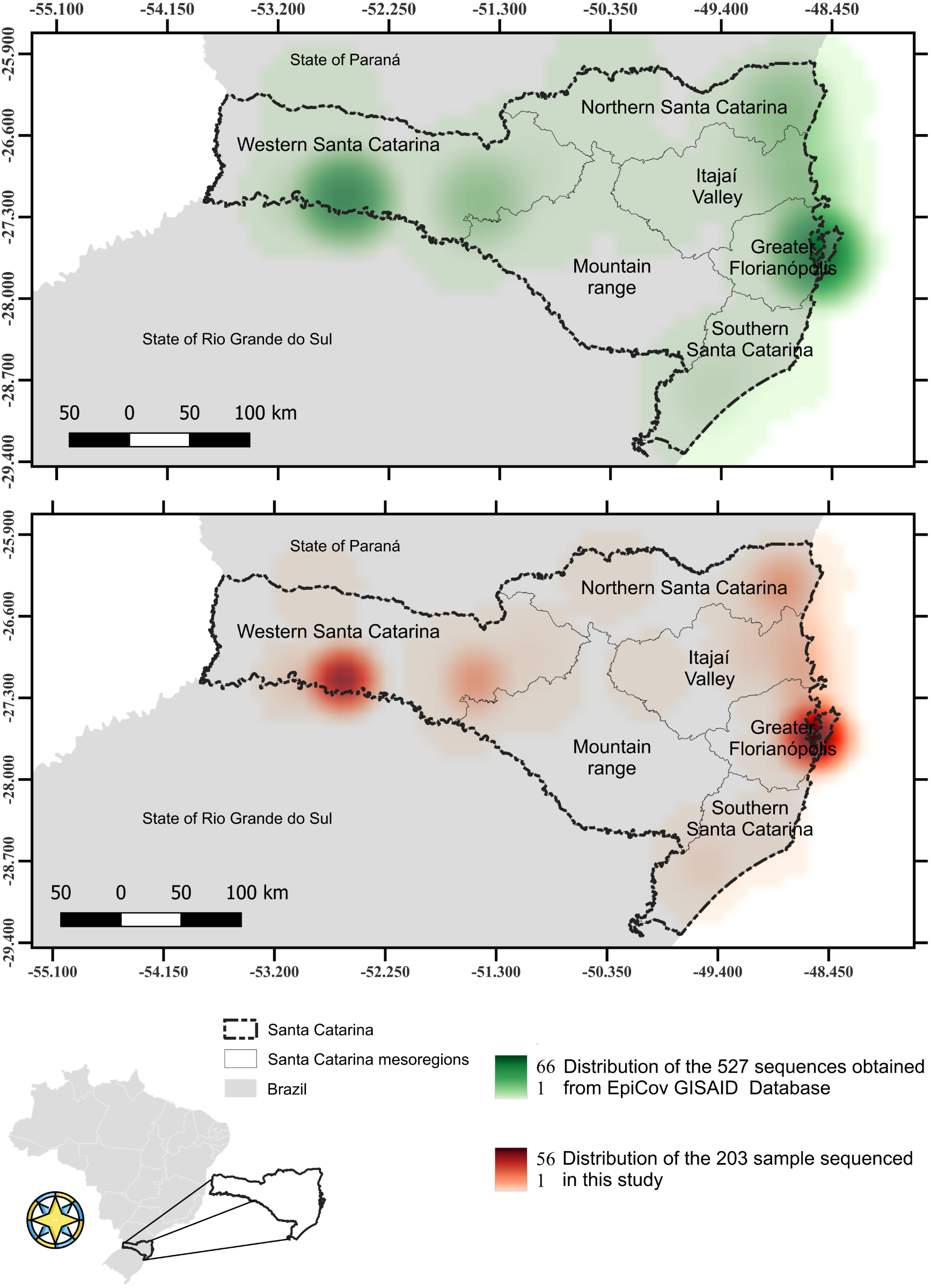
Distribution of sequenced SARS-CoV-2 genomes in Santa Catarina from March 1^st^, 2020 to April 30^th^ 2021. Map showing the Santa Catarina SARS-CoV-2 sequences obtained from GISAD EpiCov Database (upper map) and samples sequenced in this study (lower map).

### RNA purification and sequencing

The study used 203 human nasopharyngeal swab samples from SARS-CoV-2 positive patients diagnosed via RT-qPCR. Inclusion criteria was a positive RT-PCR for gene E with C_t_ values < 25. Total viral RNA was extracted using the RNA Viral Kit (QIAGEN, Hilden, Germany) and the long amplicon pooled protocol was used [17]. Briefly, complementary DNA (cDNA) was synthesized from viral RNA using 50 μM random primers and 200 U/μL Superscript IV reverse transcriptase (ThermoFisher Scientific, Waltham, MA, USA). The obtained cDNA was amplified using 10 μM of 14 primer sets in individual reactions to amplify different regions of the SARS-CoV-2 genome (approximately 2.5 Kb in each fragment) and 40 U/μL Platinum *Taq* DNA polymerase High Fidelity (Invitrogen, Waltham, MA, USA). Amplicons were visualized on agarose 2% gel electrophoresis.

The amplicons were then pooled, purified using the AMPure XP (Beckman Coulter, Brea, CA, USA) and quantified by Quant-iT Picogreen dsDNA assays (Invitrogen, Waltham, MA, USA). Pools were then processed using the Illumina DNA Prep (Illumina, San Diego, CA, USA), following manufacturer’s protocol. Libraries were quantified using the Collibri Library Quantification kit (ThermoFisher Scientific, Waltham, MA, USA), adjusted to 11.5 pM and prepared for paired-end sequencing (2x 150 bp) using the MiSeq Reagent Kit V2 (300-cycles) (Illumina, San Diego, CA, USA). Sequencing was performed on the Illumina MiSeq platform (Illumina, San Diego, CA, USA).

### SARS-CoV-2 genome assembling and variant analysis

The SARS-CoV-2 genome was assembled according to ARTIC protocol [*18*]. Briefly, low-quality bases (Q < 25) and Illumina adapters were trimmed [*19*], and mapped to the human reference genome (hg19 - GCF_000001405.13) to remove contaminants using BWA v. 0.7.17 [*20*]. Next, human-free reads were aligned to the SARS-CoV-2 reference genome (NC_045512.2) using BWA with default parameters. Ivar v. 1.3 [*21*] and mpileup function from samtools v. 1.7 [*22*] were used to perform variant calling and to generate genomes consensus sequences. Assembled genomes were analyzed in the Nextclade [*23*] and Pangolin [*24*] platforms to classify the SARS-CoV-2 variants.

Mutations, deletions, and insertions were used to assess the dissimilarity of the genomes using the vegan R package v. 2.5.6 [*25*] with the Jaccard index method. The multidimensional scaling (MDS) and the distribution of variants in SC were plotted using the ggplot2 R package v. 3.3.0 [*26*].

### Maximum-likelihood (ML) phylogenetic analysis

The ML phylogeny was performed using all 779 SARS-CoV-2 genome sequences (Supplementary Table 2). Sequences were aligned by MAFFT v. 7.310 [*27*] with default parameters and were manually curated using AliView v.1.26 [*28*]. A ML tree was constructed using the alignment, with GTR+F+I as the substitution model predicted by ModelFinder [*29*], 1,000 bootstrap runs using UFBoot tree optimization [30] and SH-like approximate likelihood ratio test (SH-aLRT) [*31*], using IQ-TREE v. 2.1.3 [*32*]. ML tree was visualized and annotated using the ggtree R package v. 2.0.4 [*33*].

### Network analysis of SC VOC Gamma and related lineages during the second wave

The network analysis was performed using 589 Brazilian VOC Gamma and related genomes retrieved from GISAID, of which, 418 were SC SAR-CoV-2 genomes from samples obtained from January 5^th^, 2021, to April 30^th^,2021, and 171 genome sequences from other Brazilian states sequenced up to January 5^th^, 2021 (Supplementary Table 3). The ML consensus tree as constructed like above, except the substitution model was GTR+F+R2. The network analysis was performed using online service StrainHub with closeness centrality [*34*], ML consensus tree, and corresponding metadata.

### Spike (S) gene sequence polymorphism and protein structure analysis

For structural analysis of S protein of VOC Gamma, 57 presented full-length S protein coverage of VOC Gamma sequenced in this study with 722 retrieved from GISAID (December 3^rd^, 2021), including the reference sequence (NC_045512.2) were aligned using MAFFT [*27*]. The mutations were represented over the 3D S protein structure PDB 6ZGG (resolution 3.8 Å). The experimental structure presents a representative coverage of the N-terminal domain (NTD) (Gln14 to Asp1146). The mutations were highlighted and colored according to observed frequencies and built with PyMOL version 2.3.3 software [*35*].

## Results

### Profile of SARS-CoV-2 variants in State of Santa Catarina after one year of COVID-19 pandemic

A total of 23 distinct SARS-CoV-2 variants were identified among the studied genomes (Figure 2A). Of these 730 complete genomes, 316 (43.2%) were classified as the VOC Gamma, followed by 175 (23.9%) of the former VOI Zeta variant (P.2) in and 69 (9.4%) of the P1-like II variant. Only four sequences of the VOC Alpha variant (B.1.1.7) were observed among the analyzed samples.

**Figure 2:**
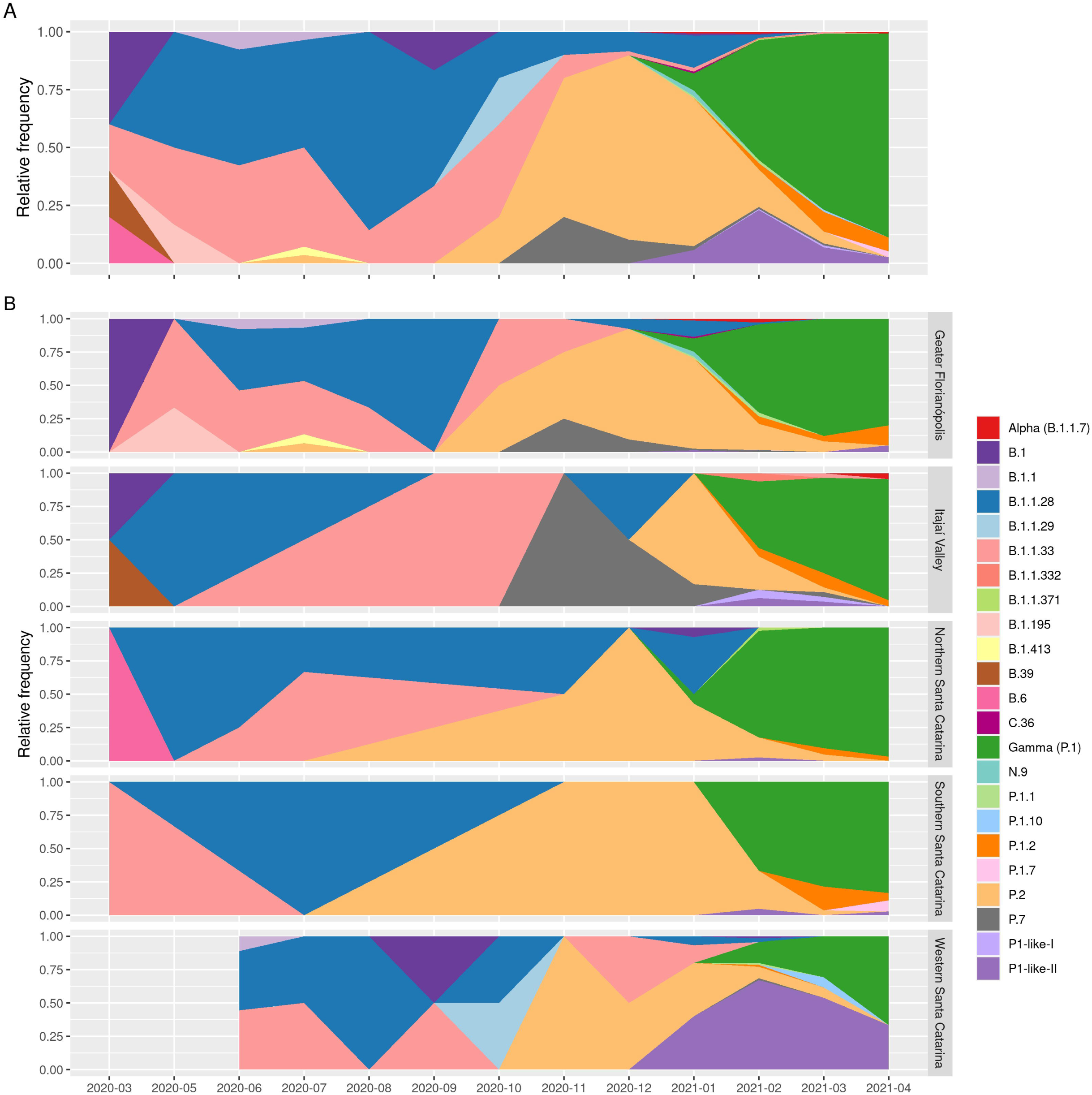
SARS-CoV-2 variants profile in the state of Santa Catarina from March 2020 to April 2021. **(A)** Analysis of the variant profile considering the whole state of Santa Catarina. **(B)** The variant profile by Santa Catarina mesoregion. From March to September 2020, the variants identified were in the highest proportion of B.1.1.28 (Blue) and B.1.1.33 (Light red). From September 2020 to January 2021, the highest proportion was of P.2 variant, followed by VOC Gamma and related lineages from February to April 2021.

The first sequenced sample was classified as a B.39 variant. Then, the B.1.1.28 and the B.1.1.33 variants prevailed from May - November 2020, when the P.2 variant gradually became the most prevalent. The P.2 variant peaked in December 2020 and was displaced quickly by the P.1 variant. By January 2021, the VOC Gamma had already taken over most of the samples in Santa Catarina (Figure 2A). Until December 2020 the profile of variants is similar among the mesoregions. However, in the beginning of the second wave (February 2021), the Western mesoregion has a distinct profile, with the highest cases/100,000 inhabitants indices (Supplementary Figure 1) and the P.1-like-II lineage being predominant in that region (Figure 2B). The Mountain Range was sampled only in February 2021, where 10 VOC Gamma and a single P.1-like-II was detected.

### VOC Gamma and related lineages in Santa Catarina

We inferred phylogenetic relationships between the SARS-CoV-2 genomes from SC and the other known variants (Figure 3). Most of the SC sequences were grouped in the VOC Gamma clade, which includes subvariants P.1.1, P.1.2 and P.1.10, and related lineages (P.1-like I and P.1-like II). However, this parent clade generates two distinct sub-clades, one containing the variant P.1-like-II and P.1-like-I, and the other clustering sequences classified as P.1 and their subvariants. It is noteworthy to mention that the clade formed mainly by P.1-like-I and P.1-like-II variants is composed of samples from the Western SC, pointing out a possible independent introduction and regionalized transmission of such variants in that region.

**Figure 3:**
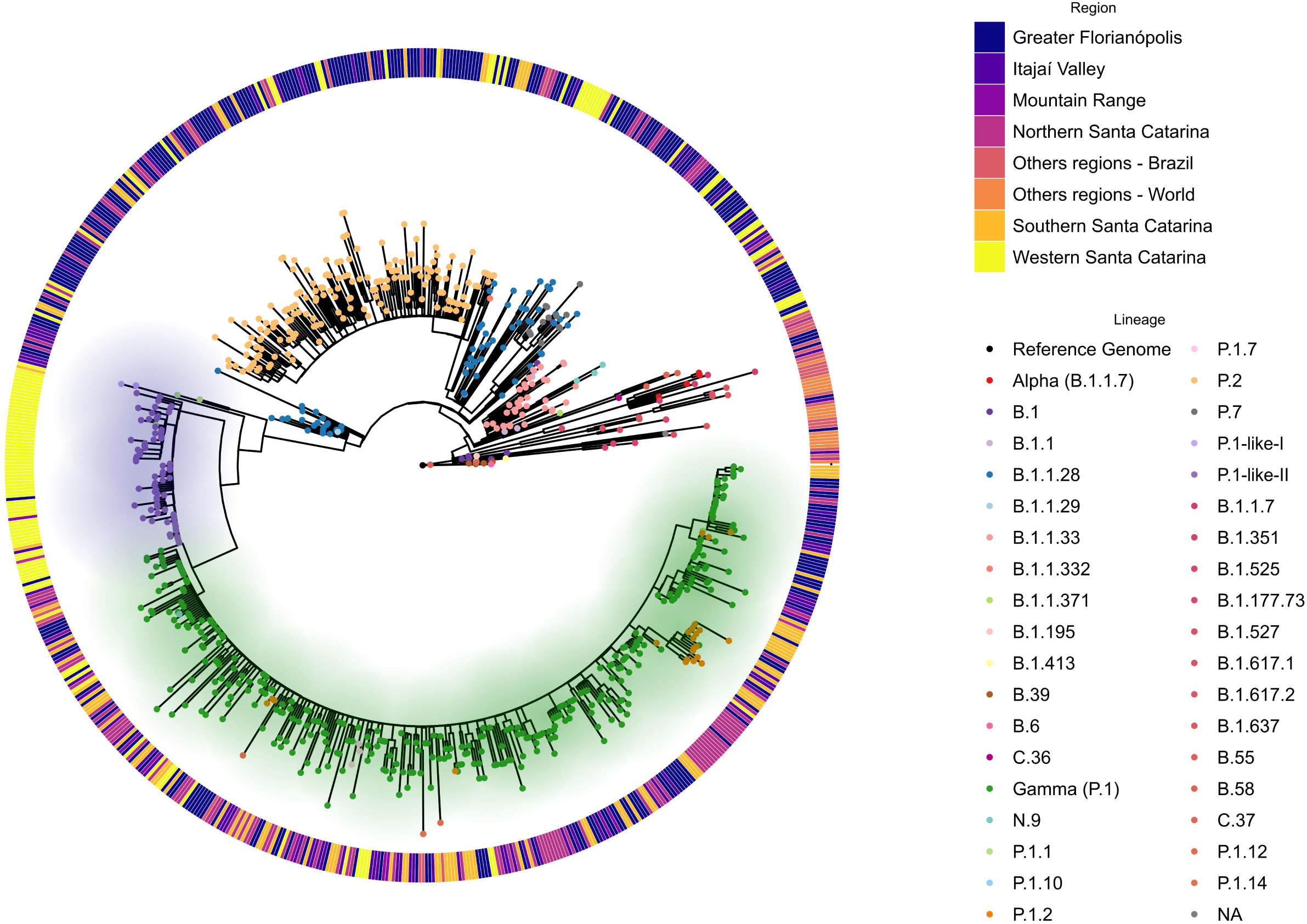
Phylogenomic reconstruction analysis of Santa Catarina SARS-CoV-2 sequences from March 1^st^, 2020 to April 30^th^ 2021. Maximum likelihood (ML) phylogenetic tree was reconstructed using GTR+F+I substitution model, with 1000 iterations in Ultra-Fast Bootstrap mode with SH-like approximate likelihood ratio test (SH-aLRT). All 779 sequences were aligned with 49 reference sequences of distinct SARS-CoV-2 variants available in GISAID. Dots represent the variant or lineages indicated on the right-bottom legend. According to the right-up legend, the outside bars indicate the sample mesoregion of Santa Catarina or others. The green shadow indicates the VOC Gamma and related lineages (P.1.1, P1.2, P.1.10 and P.1.7) clade. Lilac shadow indicates the P.1-like-II clade.

A Multidimensional scale (MDS) plot shows the clustering of the SARS-CoV-2 genomes according to the mutations observed (Figure 4A). There was no clear clustering of the studied SARS-CoV-2 variants according to the sampled geographic regions along the sampling period (Figure 4B). However, in a separate analysis of the data from the VOC Gamma and the related lineages, it was possible to observe a clustering of genomes from Western SC (Figure 4C), mainly composed by the P.1-like-II variant (Figure 4D).

**Figure 4:**
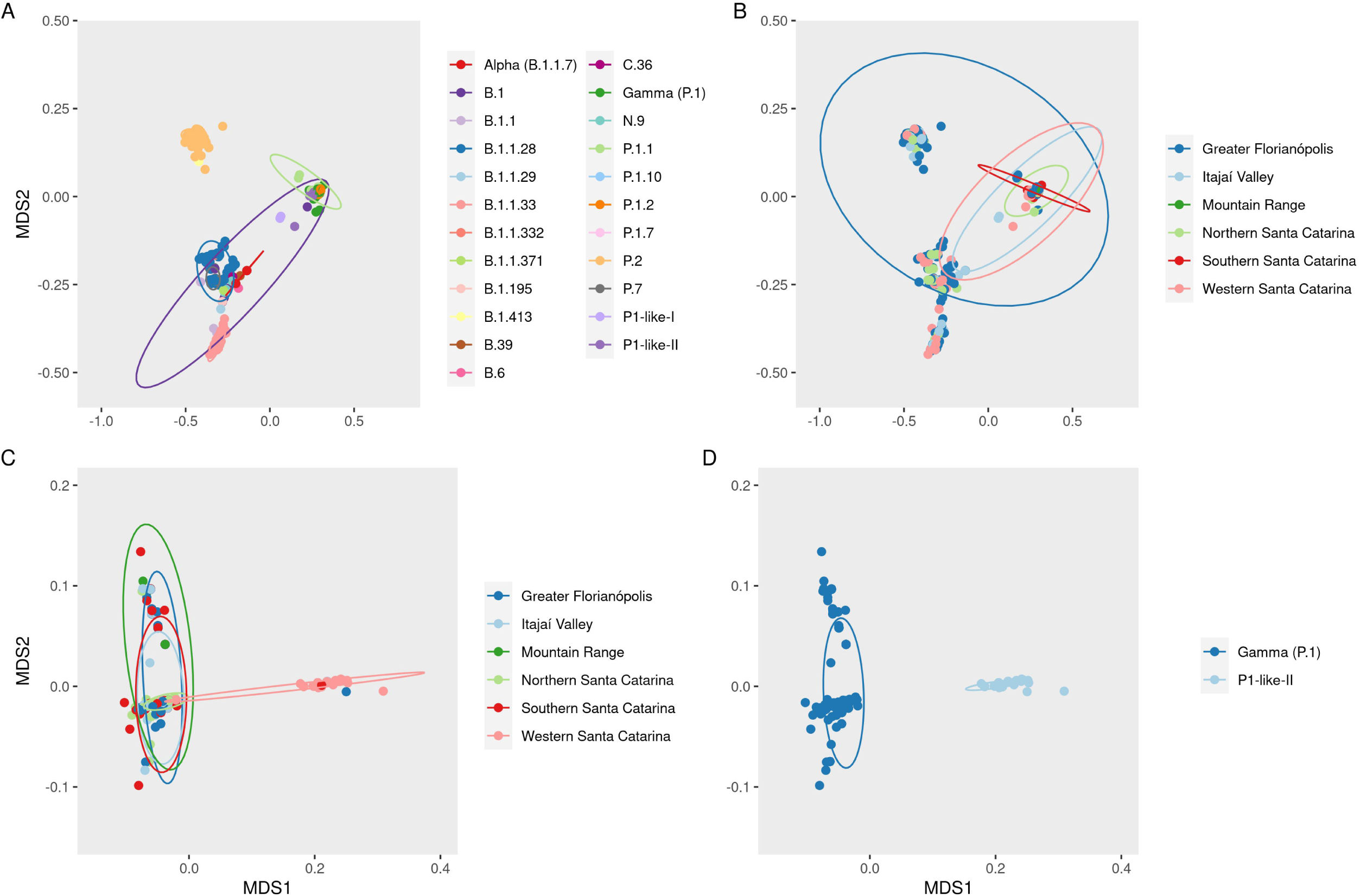
Multidimensional scale (MDS) plot of Jaccard dissimilarity matrix between genomes based on the presence and absence of mutations, deletions, and insertions labeled by **(A)** variants and **(B)** region. Only VOC Gamma and related lineages MDS analysis labeled by **(C)** region and **(D)** variant or lineages.

As the ML and the MDS analyses pointed out the evidence of genetic proximity of the cluster of P.1 related lineage (P.1-like-II) in the Western SC, we compared all SC VOC Gamma and related lineages sequences obtained from August 7^th^, 2020 to April 30^th^, 2021. These genomes were classified based on the amino acid substitutions described in P.1-like-I [*36*] and P.1-like-II [*13*] variants. Mutations that characterize each VOC Gamma related lineage profile were identified and are summarized in Supplementary Figure 2.

More than 90% of the S protein of the VOC Gamma and related lineages presents the mutation L18F, P26S, D138Y, and R190S positions of the NTD region (Figure 5). The typical mutations were found in the RBD domain (K417T, E484K, and N501Y), SD domain (D614G and H655Y), and overall structure (T1027I, and V1176F). In 20% of sequences conserved the original amino acid T20 in the NTD, being characterized as P.1-like-II lineages.

**Figure 5:**
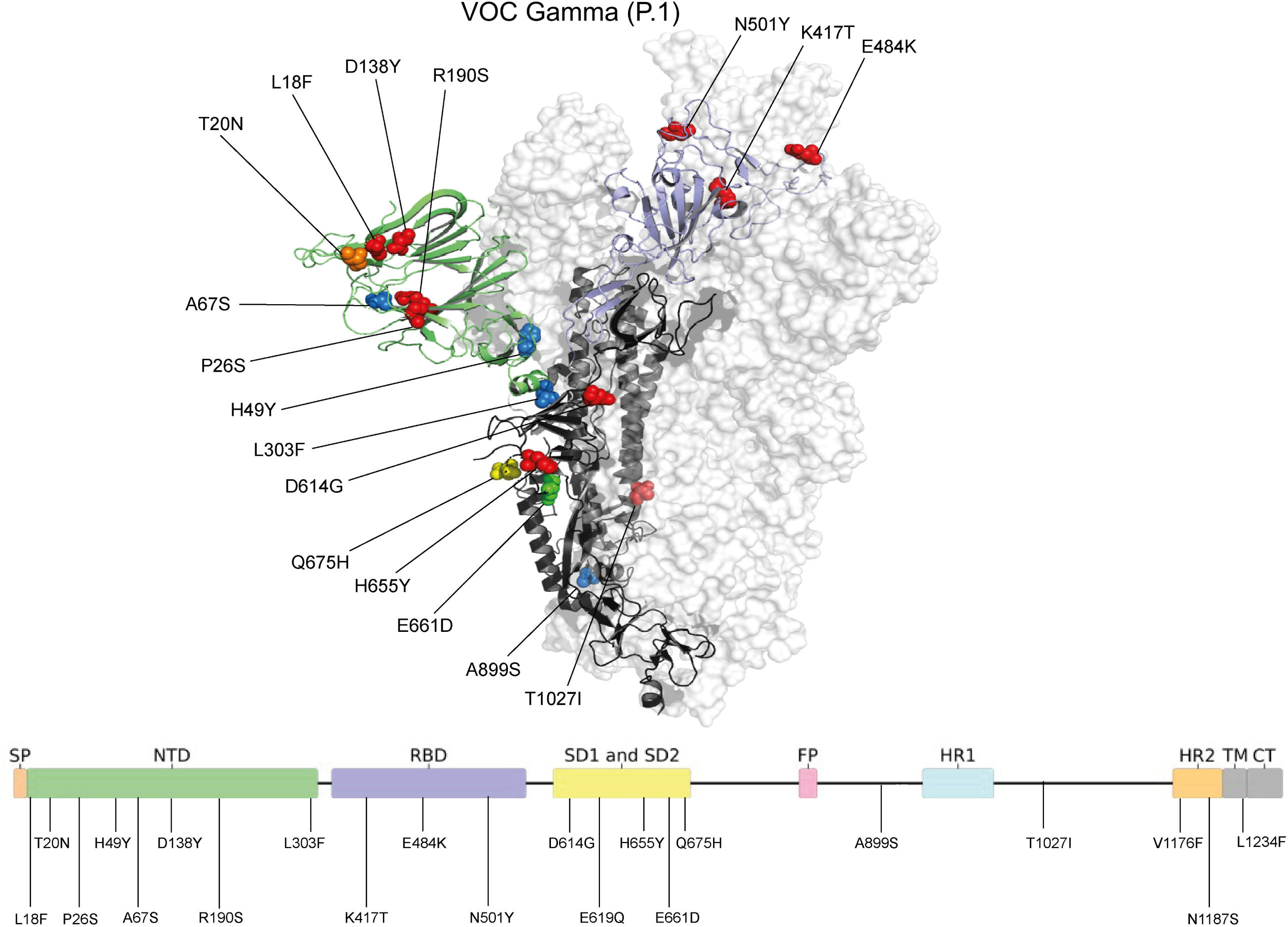
Overlay of VOC Gamma Spike protein mutations identified in the state of Santa Catarina during the COVID-19 pandemic. Residues involved in mutations are presented as spheres, colored from blue to red (low to high mutation frequency). The mutation sites are also indicated in the two-dimensional representation of the Spike protein, which is colored according to protein regions. The NTD domain is colored in green and the RBD domain is colored in purple. From the total of 778 analyzed sequences of the VOC Gamma variant (5’ – 3’), red represents the frequency of 93.6-100%, orange the frequency of 88.8%, yellow the frequency of 3.98%, green the frequency of 1.79%, and blue the frequency of less than 1%. Image generated using PyMOL version 2.3.3 [*43*].

### The rapid spread and regionalization of P.1-like-II during the second wave of COVID-19 pandemic in Santa Catarina, Brazil

During the second wave of COVID-19 (January - April 2021) the VOC Gamma and subvariants were the most representative SC. However, it does not present a homogeneous variant distribution pattern in the mesoregions (Figure 2B). Network analysis revealed that VOC Gamma and related lineage genomes had a genetic proximity with genomes sequenced in the State of Amazonas (Figure 6A). The VOC Gamma and related lineages from the Western SC had fewer interactions with other SC mesoregions than with the State of Amazonas (Figure 6B). The city of Chapecó-SC, the major municipality of the Western Santa Catarina, acted as a point of concentration and dissemination of lineages imported from Manaus to other Western cities (Figure 6C).

**Figure 6:**
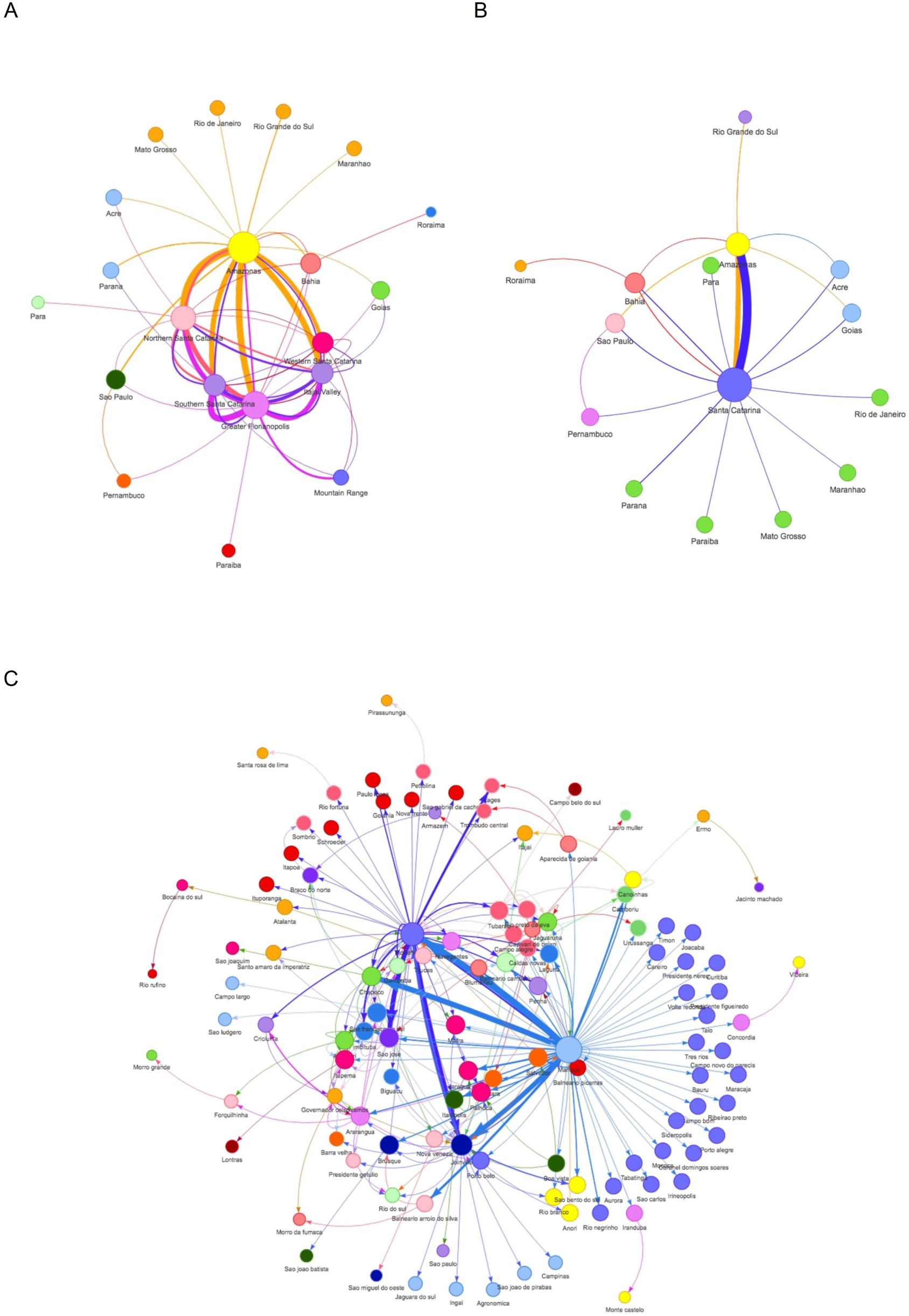
Network analysis of VOC Gamma and related lineages sequenced in the state of Santa Catarina, Brazil, from January 3^rd^, 2020 to April 30^th^ 2021 with the other Brazilian states. The ML analysis was performed on 13,018 Brazilian VOC Gamma related genomes obtained from GISAID until April 30^th^, 2021 and 316 and submitted to StrainHub [*42*] analysis considering the closeness parameter. The sizes of nodes are scaled by closeness metric and the arrows reflect the directionality of possible transmissions between the Brazilian states **(A)**, Santa Catarina mesoregions **(B)** and Santa Catarina cities **(C)**. The thickness of the lines and arrows represents the frequency of putative transmissions.

Also, it was observed that in February and March 2021, 75% and 60% of the sequences in the Western SC were classified as P.1-like-II, respectively (Figure 7). Furthermore, in the same period, this mesoregion had the highest Case Fatality Rate (CFR), mortality, and the number of deaths in the state of Santa Catarina (Figure 8), which indicates that rapid dissemination of P.1-like-II variant in the region followed by an increase in deaths.

**Figure 7:**
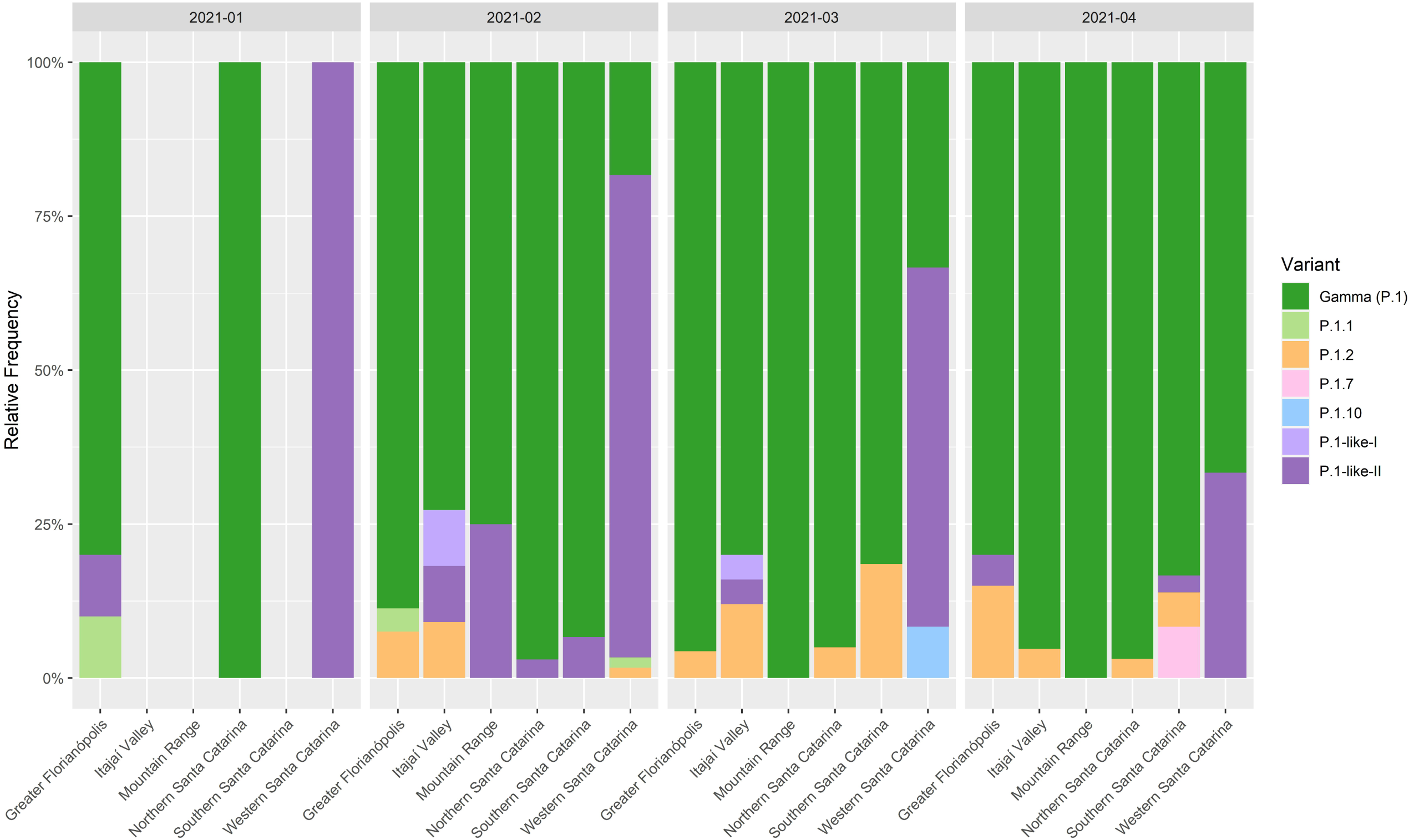
Relative frequencies of the SARS-CoV-2 VOC Gamma and related lineages by mesoregions of the state of Santa Catarina during the second wave of the COVID-19 pandemic, January to April 2021.

**Figure 8:**
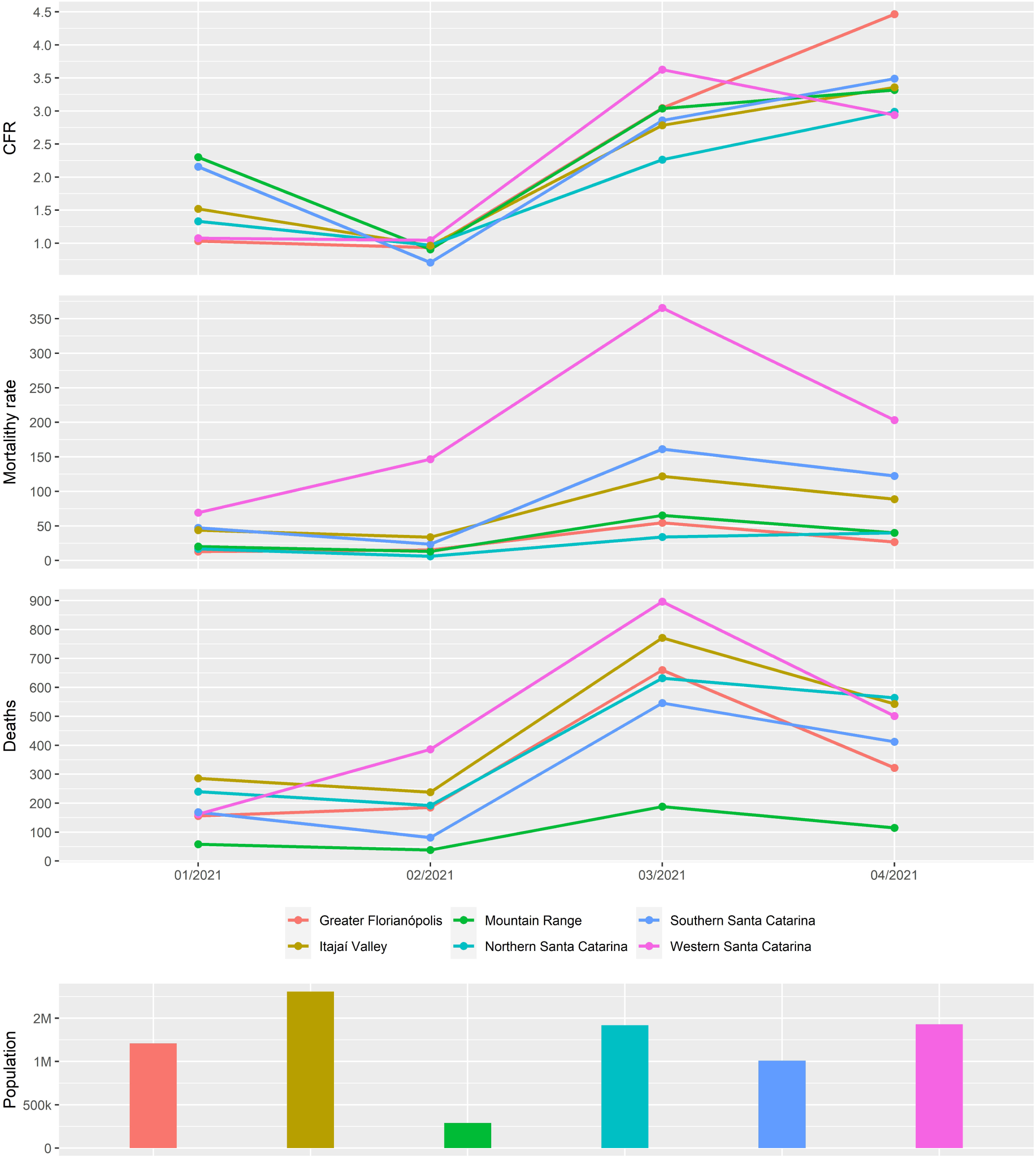
Epidemiological data from each mesoregion of the state of Santa Catarina during the second COVID-19 pandemic wave (January 2020 to April 2021). Case fatality rate (CFR), mortality rate, number of deaths and population size. Data source: Santa Catarina Epidemiological Surveillance Directorate [*16*].

Taken together, these data indicate distinct introductions of VOC Gamma and related lineages in Santa Catarina, followed by a rapid dispersion of the P.1-like-II variant in the Western region of SC and a regionalization in the transmission of P.1-like-II in the Western region.

## Discussion

In this study carried out between March 2020-April 2021, we sequenced 203 SARS-CoV-2 genomes from human samples collected from different regions of the SC state, representing 27.8% of the virus genomic sequences generated for this region. Assessment of SARS-CoV-2 genetic profiles (Figure 2A) allowed the identification of VOC Alpha (20I/501Y.V1/ B.1.1.7) and especially the Gamma (20J/501Y.V3/P.1) from January 2021 onwards, corresponding to the second wave of the pandemic in SC, as also observed in other Brazilian states [*37, 38*], indicating the homogeneity of the spreading pattern within Brazil at that time. However, between August 2020-November 2020 we were able to identify the emergence of the formerly VOI P.2 [*1*] in SC, which rapidly turned to be more prevalent than the other variants circulating in the region. This variant was originally identified in Brazil in Rio de Janeiro by July 2020, although phylogenetic analyses indicate that the variant may have arisen as early as February 2020 [*39*]. Similar to our findings in SC, P.2 was reported to have become predominant in several Brazilian states around the same period, especially in Northeastern and Southeastern Brazil, when variant P.1 turned to be more prevalent by beginning 2021 [*39*], except for the State of Amazonas where P.2 variant was first detected in November 2020 but was overshadowed by P.1 since its early detection and thus never became the main circulating variant [*38*]. After the emergence of VOC Gamma in Manaus City, State of Amazonas [*7*], the first case of P.1 SC was reported in in Florianópolis City (Greater Florianópolis) (hCoV-19/Brazil/SC-1326/2021) by January 8^th^, 2021, and the first case of VOC Gamma related lineage (P.1-like-II) in Chapecó City (Western Santa Catarina) on January 22^th^, 2021, the later also reported by Gräff et al [*12*]. The genetic variability of SARS-CoV-2 observed in our study is shown by the variants found here (Figure 2), which probably emerged because of the genome replication during the viral infection. Synthesis of RNA virus particles can produce an average of 100,000 viral copies in approximately 10 hours, with one RNA molecule produced every 0.4 seconds, without repair system of this new RNA produced [40,41] .ML analysis of P.1-like-II samples obtained from patients living in the Western mesoregion of SC were clustered in a single, separate clade (Figure 3), while viral samples from the other SC regions due to scarce P.1-like-II were mostly VOC Gamma and subvariants, pointing out that introduction and initial spreading of P.1-like-II variant in SC occurred via Western mesoregion (Figures 4C and 4D). Also, as highlighted in Figure 5 and Supplementary Figure 2, VOC Gamma is characterized by three mutations in the Receptor Binding Domain - RBD (K417T, E484K, and N501Y) that can modulate ACE2/RBD affinity, increasing transmissibility or even the affinity of antibody. In addition to mutations L18F, T20N, P26S, D138Y and R190S in the N-terminus (NTD), D614G and H655Y in the C-terminus of S1, and T1027I and V1176F in S2 are also report to VOC Gamma [*42*]. All these mutations were identified in SC, however the cluster 3 and 8 contain the P.1-like-II lineage are formed by sequences with the absence of T20N mutation and are composed specially by samples from Western region (Supplementary Figure 2). The appearance of P.1-like-II, as well as several of the variants sequenced here, is a phenomenon of cumulative mutations characteristic of RNA viruses. Accumulation of SARS-CoV-2 genome mutations may be related to transmissibility, host cell affinity, and pathogenicity [43]. These characteristics may explain what was observed in Figure 8, in which the fatalities were higher in the western region of the state of Santa Catarina, where the P.1-Like-II variant was in greater proportion.

The evolution of RNA viruses is based on amino acid substitution, the replacement rate of SARS-CoV-2 being very similar to other RNA viruses such as Influenza A and Human Enterovirus, and the replacement rate for SARS-CoV-2 is 10−3 replacement per site per year, approximately two replacements per month, for circulating viruses [44,45]. Amino acid insertions are common in Spike protein (S1 and S2), which may be a natural process of viral adaptation. Investigating changes in structural protein S is of paramount importance, given the need for this protein for virus-host cell binding [46].

Thus, in this study, analysis of such markers allowed identification of two main lineages that are distinguished by the presence or absence of the T20N mutation in the S protein. This SARS-CoV-2 protein is highly glycosylated, being involved in the virus evasion mechanisms of the human immune system through mutations that hinder viral recognition by the immune system. Most of these glycosylation sites are located in Asp residues that contains N-glycosylation sites, or Ser and Thr residues that contain O-glycosylation sites. Among these, the most frequent glycosylation sites were found in N+2, where Asp is followed by a Ser/Thr residues [*47*]. Remarkably, the herein reported T20N mutation at the NTD is followed by a Thr (N+2) in all analyzed samples (N20X21T22) and introduces a new potential O-glycosylation site within the S protein that may increase the virus capability to evade the immune defenses of viral neutralization by means of antibodies, not altering the potential N-glycosylation site at N17 that is also present in P.1 variants, where a Thr follows an Asp residue at position 19 (N17X18T19).

As shown in Figure 4D, by February 2021 we identified a high frequency of the subvariant P.1-like-II in samples from Western SC if comparing to the other regions of the state, preceding an increase in the CFR index, mortality rate, and deaths (Figure 8) on that mesoregion. Taken together, these data indicate that P.1-like-II had high transmissibility and effect on the lethality in the Western SC. Naveca et al. [*38*] showed how the second wave of COVID-19 in the state of Amazonas, with a sudden rise in cases and number of deaths during late 2020 and early 2021, directly correlates to the rise in Gamma frequency in the state, which had reached a massive prevalence by January 2021. Faria et al. [7] demonstrated that the infection by VOC Gamma is 1.1-1.8 times more severe than those previously observed by other variants, considering that the results of this variant was related to higher viral loads, greater transmissibility and an increase in the number of deaths.

The relationship between the increase in the number of cases and the appearance of variants was also observed outside of Brazil, such as in India and Europe. In India, the appearance of the Delta variant was observed during the second wave, correlating with the consequent increase in the number of deaths. In Europe, the second wave was also marked by an increase in the number of cases and deaths, which correlated with the appearance of the Alpha variant [48, 49]. However, transmission rate of SARS-CoV-2 may be affected by a variety of factors and the emergence of a given variant could not be considered the unique factor to increase such rate [*50*]. Along several virus-inherent factors such as genetic variability, the lack or misuse of non-pharmaceutical interventions (NPI), social difficulties for diagnosis or late diagnosis, uncontrolled population movement [*51*], the collapse of the public health system [*52*], lower vaccination [*53*], the overtime decrease of the immune protection [*52*], late or misdecisions taken by authorities [*61*], and the use of scientifically unproven preventive therapies [*56-58*], are also important factors that might contribute to increase the transmission and case fatality rates. Examples in different countries show that an increase in the movement of people and low widespread adherence to the NPI was observed weeks before the collapse of the public health service, and the consequent increase in the number of cases and deaths [*59-60*]. Indeed, the SARS-CoV-2 circulation is evidenced by the detection of viral RNA copies in sewage months before being reported in people, as occurred in Santa Catarina state in November 2019 [61]. Thus, we cannot rule out that conjunction of both viral and other biological, environmental and behavioral factors have significant influence on the occurrence of SARS-CoV-2 variants presenting variable infectivity and pathological effects. Such may have contributed to the differentially increased number of cases, hospitalization, and deaths (CRF) in a short period of time in the Western SC as also previously observed in Manaus (Amazon, Brazil) having totally distinct environmental conditions than SC.

With the following report of the VOC Omicron in South Africa in November 2021 [*1*] this variant was rapidly detected worldwide [1] and reported in SC, beginning of December 2021, evidencing the ongoing viral adaptations and the continuing pandemic spreading pattern. Considering these facts, it is of utmost importance the needs for maintenance of a detailed and standardized genomic surveillance of SARS-CoV-2 variants towards precise and specific identification of VOCs, avoiding erroneous epidemiological inferences and allowing evaluation of vaccine efficacy across viral variants.

Genomic surveillance of the SARS-CoV-2 pandemic has been crucial to assess the accumulation of mutations, mostly due to the enormous quantity of viral replication events, and the biological outcomes of the genetic variability of each variant on the epidemiology, transmissibility, infectivity, pathogenesis and vaccine efficacy and effectiveness. Since major concerns over the genetic evolution of SARS-CoV-2 are related to an increased spreading capability and to the escape from the immune system, genomic surveillance provides essential data to prevent the current pandemic from lasting even longer.

## Conclusion

This study allowed us to observe the distinct patterns of circulation of different SARS-CoV-2 variants within SC regions. Our data suggest distinct introduction, and rapid spreading of the P.1-like-II in the Western regions of SC in the initial months of 2021, where it was responsible for distinct and increased hospitalization and mortality rates. Thus, the continuous real-time genomic surveillance of SARS-CoV-2 in Santa Catarina can offer precise epidemiological support for the implementation and review of policies and actions carried out by public health authorities towards control of COVID-19 transmission and assessment of immunization efficacy.

## Data Availability

All data produced in the present work are contained in the manuscript

## Acknowledgments

This work is dedicated to all SC citizens who suffered or passed away due to COVID-19. We are indebted to Hospital Universitário/HU/EBSERH from Federal University of Santa Catarina (Florianópolis, SC, Brazil), Hospital Regional do Oeste (Chapecó, SC, Brazil) and Hospital Regional Santa Terezinha (Joaçaba, SC, Brazil) for granting access to interview patients and perform sample collection. We also are thankful to all institutions that provide access to genome data at EpiCov (GISAID). Finally, we are thankful to SeTIC/UFSC (Superintendence of Electronic Governance and Information and Communication Technology) for the computational infrastructure support.

## Disclaimers

The opinions expressed by authors contributing to this journal do not necessarily reflect the opinions of the Federal University of Santa Catarina, Brazil, or the institutions with which the authors are affiliated. Funders have no role on study design, data analysis or decision to publish.

**Author Bio** (first author only, unless there are only 2 authors)

Dayane A. Padilha is a PhD student in Biotechnology and Biosciences at the Federal University of Santa Catarina (UFSC) and recipient of a CAPES scholarship. She works at both Applied Virology and Bioinformatics Laboratories at UFSC. Her main interest is on virus genomics applied to genomic surveillance.

## Supplemental Materials

**Supplementary Table 1:** Synthesis of 203 samples sequenced in the present study. Sequence name, GISAD accession numbers, city, mesoregion, age, gender, variant and clade assignment.

**Supplementary Table 2:** List of GISAD accession numbers of sequences used on ML analysis, by Pangolin lineage.

**Supplementary Table 3:** GISAD accession numbers of sequences used on Santa Catarina VOC Gamma and related lineages network analysis.

**Supplementary Figure 1:** Spatial and temporal evolution of COVID-19 cases in the state of Santa Catarina, according to the Santa Catarina Epidemiological Surveillance Directorate [*16*] during the study period. The Greater Florianópolis and the Western Santa Catarina mesoregions were the most represented because they presented the highest cases / 100,000 habitants rates between January 1^st^ 2021 and April 30^th^ 2021.

**Supplementary Figure 2:** The non-synonymous amino acid substitution profile of the VOC Gamma and related lineages genomes in the state of Santa Catarina from January 3^rd^ 2020 to April 30^th^ 2021.

## References

1. World Health Organization. WHO Coronavirus (COVID-19) [Internet]. 2019 Dec 30 [cited 2021 Dec 12] https://www.who.int

2. National Council of Health Secretaries - Covid-19 CONASS Panel [Internet]. 2021 May [cited 2021 Dec 4] https://www.conass.org.br/painelconasscovid19/.

3. Singh J, Pandit P, McArthur AG, Banerjee A, Mossman K. Evolutionary trajectory of SARS-CoV-2 and emerging variants. Virol J. 2021 Aug 13;18(1):166.

4. Meredith LW, Hamilton WL, Warne B, Houldcroft CJ, Hosmillo M, Jahun AS, et al. Rapid implementation of SARS-CoV-2 sequencing to investigate cases of healthcare associated COVID-19: a prospective genomic surveillance study. Lancet Infect Dis. 2021 Mar;21(3):e36. Corrected and republished from Lancet Infect Dis. 2020 Nov;20(11):1263–1272.

5. Global Initiative on Sharing Avian Influenza Data. [Internet]. 2008 [cited 2021 Nov 27] https://www.gisaid.org.

6. Tao, K., Tzou, P.L., Nouhin, J. et al. The biological and clinical significance of emerging SARS-CoV-2 variants. Nat Rev Genet. 2021; 22: 757–773.

7. Faria NR, Mellan TA, Whittaker C, Claro IM, Candido DDS, Mishra S, et al. Genomics and epidemiology of P.1 SARS-CoV-2 lineage in Manaus, Brazil. Science. 2021 May 21; 372(6544):815–821.

8. Hemmer CJ, Löbermann M, Reisinger EC. COVID-19: epidemiology and mutations: An update [in German]. Radiologe. 2021 Oct;61(10):880–887.

9. Nonaka CKV, Gräf T, Barcia CAL, Costa VF, de Oliveira JL, Passos RDH, et al. SARS-CoV-2 variant of concern P.1 (Gamma) infection in young and middle-aged patients admitted to the intensive care units of a single hospital in Salvador, Northeast Brazil, February 2021. Int J Infect Dis. 2021 Oct;111:47–54.

10. Greaney AJ, Starr TN, Gilchuk P, Zost SJ, Binshtein E, Loes AN, et al. Complete Mapping of Mutations to the SARS-CoV-2 Spike Receptor-Binding Domain that Escape Antibody Recognition. Cell Host Microbe. 2021 Jan 13;29(1):44–57.

11. Liu Z, VanBlargan LA, Bloyet LM, Rothlauf PW, Chen RE, Stumpf S, Zhao H, Errico JM, Theel ES, Liebeskind MJ, Alford B, Buchser WJ, et al. Identification of SARS-CoV-2 spike mutations that attenuate monoclonal and serum antibody neutralization. Cell Host Microbe. 2021 Mar 10;29(3):477–488.e4. doi: 10.1016/j.chom.2021.01.014.

12. Gräf T, Bello G, Venas TMM, Pereira EC, Paixão ACD, Appolinario LR, et al. Identification of SARS-CoV-2 P.1-related lineages in Brazil provides new insights about the mechanisms of emergence of Variants of Concern. [Internet]. 2021 Jun [cited 2021 Nov 30] https://doi.org/10.21203/rs.3.rs-580195/v1.

13. Varela APM, Prichula J, Mayer FQ, Salvato RS, Sant’Anna FH, Gregianini TS, Martins LG, Seixas A, Veiga ABGD. SARS-CoV-2 introduction and lineage dynamics across three epidemic peaks in Southern Brazil: massive spread of P.1. Infect Genet Evol. 2021 Dec;96:105144.

14. Oliveira MM, Schemberger MO, Suzukawa AA, Riediger IN, do Carmo Debur M, Becker G, et al. Re-emergence of Gamma-like-II and emergence of Gamma-S:E661D SARS-CoV-2 lineages in the south of Brazil after the 2021 outbreak. Virol J. 2021 Nov 17;18(1):222.

15. Brazilian Institute of Geography and Statistics - Official Territorial Area - Federative Unit Consultation [Internet]. 2010 [cited 2021 Oct 15] https://www.ibge.gov.br/cidades-e-estados/sc.html.

16. Santa Catarina Epidemiological Surveillance Directorate [Internet]. 2020 Mar [cited 2021 Nov 7] https://www.dive.sc.gov.br/.

17. Eden J-S., Sim E. SARS-CoV-2 Genome Sequencing Using Long Pooled Amplicons on Illumina Platforms. [Internet]. 2020 Apr 04 [cited 2021 Nov 24] https://www.protocols.io/view/sars-cov-2-genome-sequencing-using-long-pooled-amp-befyjbpw?step=81.

18. Young, E., Oakeson, K. Utah DoH ARTIC/Illumina Bioinformatic Workflow [Internet]. 2021 Jan [cited 2021 Nov 28] https://github.com/CDCgov/SARS-CoV-2_Sequencing/tree/master/protocols/BFX-UT_ARTIC_Illumina.

19. Bolger, A. M., Lohse, M., Usadel, B. Trimmomatic: a flexible trimmer for Illumina sequence data. Bioinformatics. 2014 Aug 1;30(15):2114–20.

20. Li H, Durbin R. Fast and accurate short read alignment with Burrows-Wheeler transform. Bioinformatics. 2009 Jul 15;25(14):1754–60.

21. Grubaugh ND, Gangavarapu K, Quick J, Matteson NL, De Jesus JG, Main BJ, et al. An amplicon-based sequencing framework for accurately measuring intrahost virus diversity using PrimalSeq and iVar. Genome Biol. 2019 Jan 8;20(1):8.

22. Danecek, P. et al. Twelve years of SAMtools and BCFtools. Gigascience. 2021 Feb 16;10(2):giab008.

23. Nextclade Web 1.10.0 [Internet]. 2020 Jun 11 [cited 2021 Nov 9]. https://clades.nextstrain.org/.

24. Pangolin COVID-19 Lineage Assigner [Internet]. 2021 [cited 2021 Nov 9]. https://pangolin.cog-uk.io/.

25. Oksanen J, Blanchet FG, Friendly M, Kindt R, Legendre P, McGlinn D, et al. Vegan: Community Ecology Package. R package version 2.0-2 [Internet]. 2020 Nov 28 [cited 2021 Dec 5] https://CRAN.R-project.org/package=vegan.

26. Wickham, H. Ggplot2: Elegant Graphics for Data Analysis (ed. Springer-Verlag) cheatsheet. New York; 2016.

27. Katoh K, Standley DM. A simple method to control over-alignment in the MAFFT multiple sequence alignment program. Bioinformatics. 2016 Jul 1;32(13):1933–42.

28. Larsson A. AliView: a fast and lightweight alignment viewer and editor for large datasets. Bioinformatics. 2014 Nov 15;30(22):3276–8.

29. Kalyaanamoorthy S, Minh BQ, Wong TKF, von Haeseler A, Jermiin LS. ModelFinder: fast model selection for accurate phylogenetic estimates. Nat Methods. 2017 Jun;14(6):587–589.

30. Hoang DT, Chernomor O, von Haeseler A, Minh BQ, Vinh LS. UFBoot2: Improving the Ultrafast Bootstrap Approximation, Molecular Biol. Evol. 2018 Feb;35(2):518–522.

31. Guindon S, Dufayard J-F, Lefort V, Anisimova M, Hordijk W, Gascuel O. New Algorithms and Methods to Estimate Maximum-Likelihood Phylogenies: Assessing the Performance of PhyML 3.0, System. Biol. 2010 May;59(3):307–321.

32. Minh BQ, Schmidt HA, Chernomor O, Schrempf D, Woodhams MD, von Haeseler A, et al. IQ-TREE 2: New Models and Efficient Methods for Phylogenetic Inference in the Genomic Era. Mol Biol Evol. 2020 May 1;37(5):1530–1534.

33. Yu G. Using ggtree to Visualize Data on Tree-Like Structures. Curr Protoc Bioinformatics. 2020 Mar;69(1):e96.

34. Schneider AB, Ford CT, Hostager R, Williams J, Cioce M, Çatalyürek ÜV, et al. StrainHub: a phylogenetic tool to construct pathogen transmission networks. Bioinformatics. 2020 Feb 1;36(3):945–947.

35. Schrödinger, LLC. The PyMOL Molecular Graphics System, Version 2.0 [Internet]. 2017 Sep 20 [cited 2021 Nov 17] https://pymol.org/2/.

36. Naveca F, Nascimento V, Souza V, Corado A, Nascimento F, Silva G, et al. Phylogenetic relationship of SARS-CoV-2 sequences from Amazonas with emerging Brazilian variants harboring mutations E484K and N501Y in the Spike protein. Genom. Epidemiol. [Internet]. 2021 Jan 11 [cited 2021 Nov 12] https://virological.org/t/phylogenetic-relationship-of-sars-cov-2-sequences-from-amazonas-with-emerging-brazilian-variants-harboring-mutations-e484k-and-n501y-in-the-spike-protein/585.

37. Castro, M. C. et al. Spatiotemporal pattern of COVID-19 spread in Brazil. Science. 2021 May 21;372(6544):821–826.

38. Naveca FG, Nascimento V, de Souza VC, Corado AL, Nascimento F, Silva G, et al. COVID-19 in Amazonas, Brazil, was driven by the persistence of endemic lineages and P.1 emergence. Nat Med. 2021 Jul;27(7):1230–1238.

39. Lamarca AP, de Almeida LGP, Francisco RDS Jr, Lima LFA, Scortecci KC, Perez VP, et al. Genomic surveillance of SARS-CoV-2 tracks early interstate transmission of P.1 lineage and diversification within P.2 clade in Brazil. PLoS Negl Trop Dis. 2021 Oct 13;15(10):e0009835.

40. Domingo E, Holland JJ. RNA virus mutations and fitness for survival. Annu Rev Microbiol. 1997;51:151–78.

41. Drake JW, Holland JJ. Mutation rates among RNA viruses. Proc Natl Acad Sci U S A 1999 Nov 23;96(24):13910–3.

42. Dejnirattisai, W et al. Antibody evasion by the P.1 strain of SARS-CoV-2. Cell. 2021 May 27;184(11):2939–2954.e9.

43. Giovanetti M, Benedetti F, Campisi G, Ciccozzi A, Fabris S, Ceccarelli G, Tambone V, Caruso A, Angeletti S, Zella D, Ciccozzi M. Evolution patterns of SARS-CoV-2: Snapshot on its genome variants. Biochemical and Biophysical Research Communications. 2021; Volume 538, 2021, Pages 88–91.

44. Duchene S, Featherstone L, Haritopoulou-Sinanidou M, Rambau At, Lemey P, Baele G. Temporal signal and the phylodynamic threshold of SARS-CoV-2. Virus Evolution. 2020; Volume 6, Issue 2, veaa061.

45. Van Dorp L, Acman M, Richard D, Shaw LP, Ford CE, Ormond L, Owen CJ, Pang J, Tan CCS, Boshier FAT, Ortiz AT, Balloux F. Emergence of genomic diversity and recurrent mutations in SARS-CoV-2. Infection, Genetics and Evolution. 2020; Volume 83, 104351. ISSN 1567-1348.

46. Zhou P, Yang XL, Wang XG, Hu B, Zhang L, Zhang W, Si HR, Zhu Y, Li B, Huang CL, Chen HD, Chen J, Luo Y, Guo H, Jiang RD, Liu MQ, Chen Y, Shen XR, Wang X, Zheng XS, Zhao K, Chen QJ, Deng F, Liu LL, Yan B, Zhan FX, Wang YY, Xiao GF, Shi ZL. A pneumonia outbreak associated with a new coronavirus of probable bat origin. Nature. 2020; 579, 270–273.

47. Tian W, Li D, Zhang N, Bai G, Yuan K, Xiao H, et al. O-glycosylation pattern of the SARS-CoV-2 spike protein reveals an “O-Follow-N” rule. Cell Res. 2021 Oct;31(10):1123–1125.

48. Arnab Sarkar, Alok Kumar Chakrabarti, Shanta Dutta. Pathogens. Covid-19. Infection in India: A Comparative Analysis of the Second Wave with the First Wave. Pathogens. 2021 Sep 21;10(9):1222.

49. Katarzyna J, Samuel A, Pascal A and Mondher T. On the association between SARS-COV-2 variants and COVID-19 mortality during the second wave of the pandemic in Europe. Journal of Market Access & Health Policy. 2021 VOL. 9, 2002008.

50. Sabino EC, Buss LF, Carvalho MPS, Prete CA Jr, Crispim MAE, Fraiji NA, et al. Resurgence of COVID-19 in Manaus, Brazil, despite high seroprevalence. Lancet. 2021 Feb 6;397(10273):452–455.

51. Flaxman S, Mishra S, Gandy A, Unwin HJT, Mellan TA, Coupland H, et al. Estimating the effects of non-pharmaceutical interventions on COVID-19 in Europe. Nature. 2020 Aug;584(7820):257–261.

52. da Silva SJR, Pena L. Collapse of the public health system and the emergence of new variants during the second wave of the COVID-19 pandemic in Brazil. One Health. 2021 Dec;13:100287.

53. Kupek E. Low COVID-19 vaccination coverage and high COVID-19 mortality rates in Brazilian elderly. Rev Bras Epidemiol. 2021 Sep 10;24:e210041.

54. Lumley SF, O’Donnell D, Stoesser NE, Matthews PC, Howarth A, Hatch SB, et al. Antibody Status and Incidence of SARS-CoV-2 Infection in Health Care Workers. N Engl J Med. 2021 Feb 11;384(6):533–540.

55. Malta M, Strathdee SA, Garcia PJ. The Brazilian tragedy: Where patients living at the ‘Earth’s lungs’ die of asphyxia, and the fallacy of herd immunity is killing people. EClinicalMedicine. 2021 Feb 12;32:100757.

56. Furlan L, Caramelli B. The regrettable story of the “Covid Kit” and the “Early Treatment of Covid-19” in Brazil. Lancet Reg Health Am. 2021 Dec;4:100089.

57. Abella BS, Jolkovsky EL, Biney BT, Uspal JE, Hyman MC, Frank I, et al. Efficacy and Safety of Hydroxychloroquine vs Placebo for Pre-exposure SARS-CoV-2 Prophylaxis Among Health Care Workers - A Randomized Clinical Trial. JAMA Intern Med. 2021 Sep 30;181(2):195–202.

58. Popp M, Stegemann M, Metzendorf MI, Gould S, Kranke P, Meybohm P, Skoetz N, Weibel S. Ivermectin for preventing and treating COVID-19. Cochrane Database Syst Rev. 2021 Jul 28;7(7):CD015017.

59. Garcia LP, Duarte E. Nonpharmaceutical interventions for tackling the COVID-19 epidemic in Brazil. Epidemiol Serv Saude. 2020 Apr 9;29(2):e2020222.

60. Djin-Ye O, Silke B, Barbara B, Janine R, Frank S, Susanne D, Marianne W, Max VK, M., Thorsten W, Ralf D. Trends in respiratory virus circulation following COVID-19-targeted nonpharmaceutical interventions in Germany, January - September 2020: Analysis of national surveillance data. Lancet Reg Health Eur. 2021 Jul;6:100112.

61. Fongaro G, Stoco PH, Souza DSM, Grisard EC, Magri ME, Rogovski P, Schörner MA, Barazzetti FH, Christoff AP, de Oliveira LFV, Bazzo ML, Wagner G, Hernández M, Rodríguez-Lázaro D. The presence of SARS-CoV-2 RNA in human sewage in Santa Catarina, Brazil, November 2019. Sci Total Environ. 2021 Jul 15;778:146198.

