## Supplementary figures and images for "Emergence of two distinct SARS-CoV-2 Gamma variants and the rapid spread of P.1-like-II SARS-CoV-2 during the second wave of COVID-19 in Santa Catarina, Southern Brazil"

### Supplementary Figure 1

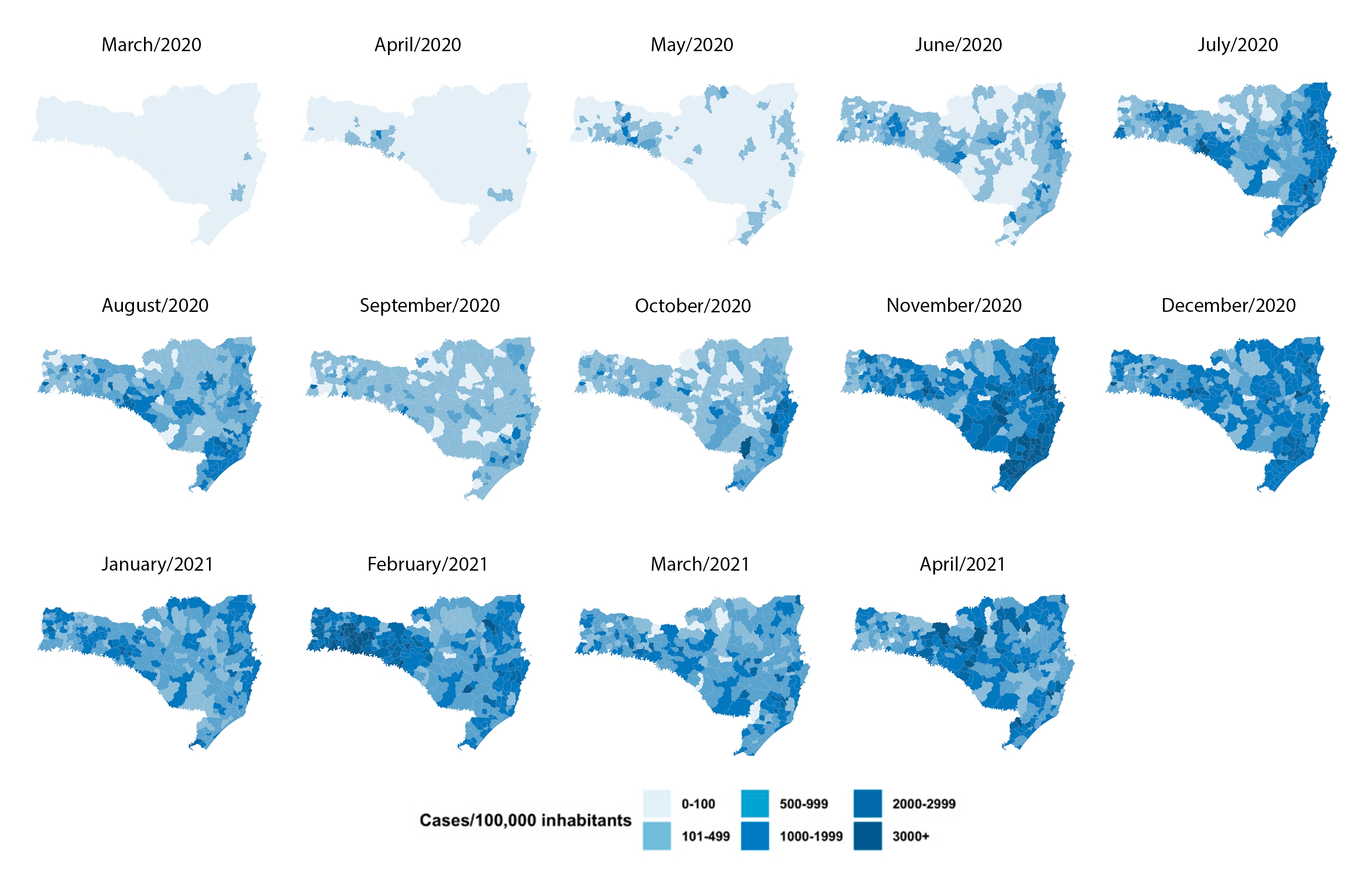

### Supplementary Figure 2

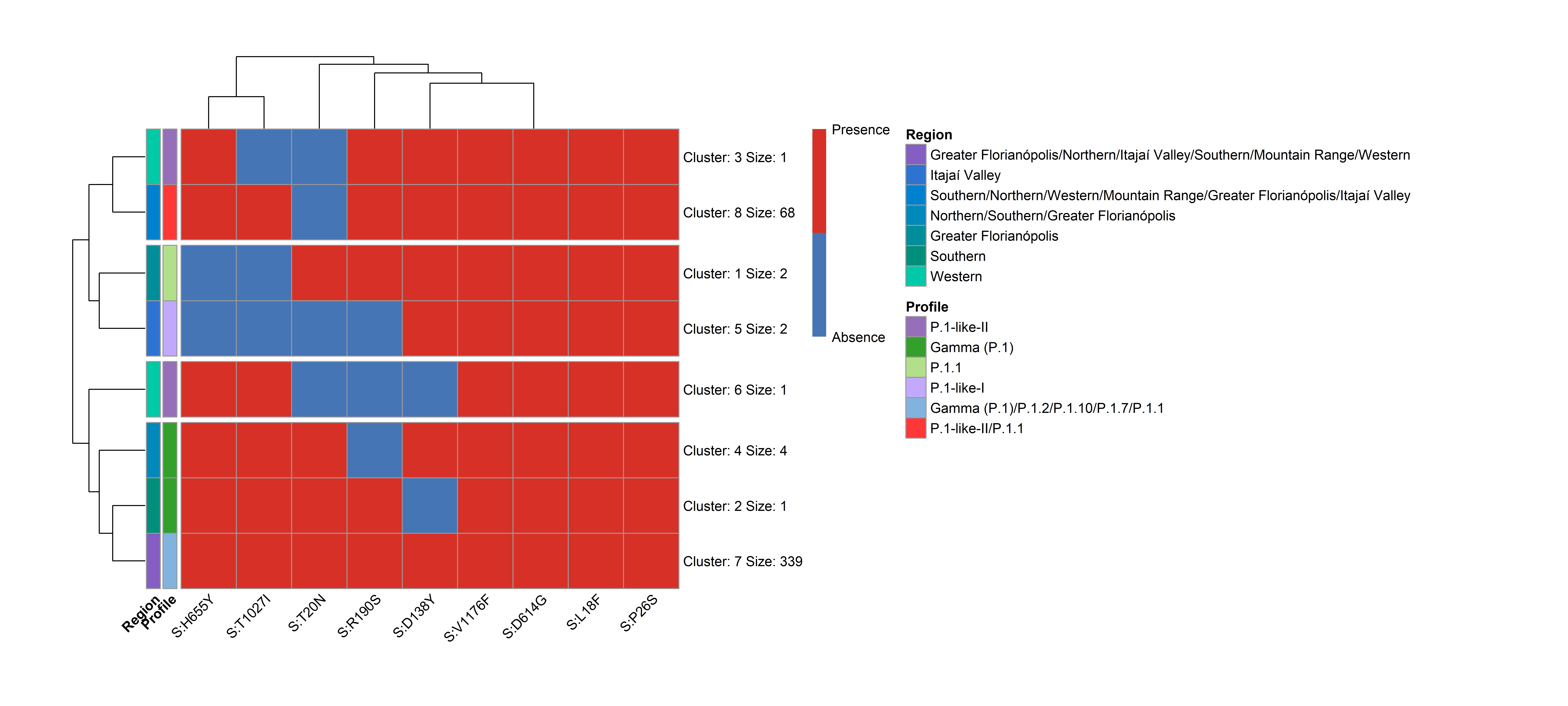
